# COVID-19 mortality across selected states in India: the role of age structure

**DOI:** 10.1101/2021.07.18.21259093

**Authors:** Mohamed Jainul Azarudeen, Khyati Aroskar, Karishma Krishna Kurup, Tanzin Dikid, Himanshu Chauhan, S.K. Jain, S.K. Singh

## Abstract

**Background:** Mortality rates provide an opportunity to identify and act on the health system intervention for preventing deaths. Hence, it is essential to appreciate the influence of age structure while reporting mortality for a better summary of the magnitude of the epidemic.

**Objectives:** We described and compared the pattern of COVID-19 mortality standardized by age between selected states and India from January to November 2020.

**Methods:** We initially estimated the Indian population for 2020 using the decadal growth rate from the previous census (2011). This was followed by estimations of crude and age-adjusted mortality rate per million for India and the selected states. We used this information to perform indirect standardization and derive the age-standardized mortality rates for the states for comparison. In addition, we derived a ratio for age-standardized mortality to compare across age groups within the state. We extracted information regarding COVID-19 deaths from the Integrated Disease Surveillance Programme special surveillance portal up to November 16, 2020.

**Results:** The crude mortality rate of India stands at 88.9 per million population(118,883/1,337,328,910). Age-adjusted mortality rate (per million) was highest for Delhi (300.5) and lowest for Kerala (35.9).The age-standardized mortality rate (per million) for India is (<15 years=1.6, 15–29 years=6.3, 30–44 years=35.9, 45– 59 years=198.8, 60–74 years=571.2, ≥75 years=931.6). The ratios for age-standardized mortality increase proportionately from 45-59 years age group across all the states.

**Conclusion:** There is high COVID-19 mortality not only among the elderly ages, but we also identified heavy impact of COVID-19 on the working population. Therefore, we recommend further evaluation of age-adjusted mortality for all States and inclusion of variables like gender, socio-economic status for standardization while identifying at-risk populations and implementing priority public health actions.

## Introduction

World Health Organization declared COVID-19 a pandemic on March 11, 2020.^1^ In India, the first case was reported on January 27, 2020, in a 20-year-old female at Thrissur city, Kerala. As of early November 2020, India recorded the second biggest number of Covid-19 cases (8 million), next only to the US (9.2 million). India is third in the number of deaths (121,144), behind Brazil (159,033) and the US (234,222). Given the population of India, these numbers may seem comparable, but most numbers are concentrated in specific states of India.

Mortality rates provide an opportunity to identify and act on the health system intervention for preventing deaths.^2^ The Sustainable Development Goal-3 uses mortality as a marker to gauge and recognize preventable deaths to fortify early recognition and hazard reduction interventions.^3^ COVID-19 mortality is an important estimate to know the disease burden in the community.^4^ Case fatality rate (CFR), the ratio of COVID-19 deaths to diagnosed cases, is the commonly reported measure to quantify mortality due to a disease. This measure is dependent on reported case numbers which can vary due to the fluctuations in testing protocols, changing case definitions, and surveillance capacity.^5^ This poses a dilemma in a populous country like India, which houses different age structures in its states and has reportedly significant difference in adult mortality within districts.^6^ The cases may be missed, but deaths are tangible. Thus, the regional disparity needs to be studied to assess the COVID-19 pandemic situation in India better. Hence it is necessary to appreciate the influence of age structure while reporting mortality for a better summary of the magnitude of the epidemic.^7^

With this objective in mind, we examined the pattern of COVID-19 mortality of India and selected states, taking into account the respective age structure and presenting the crude, age-adjusted mortality rate and age-standardized mortality rates from January to November 2020.

## Methodology

We reviewed the available reports of all the states for the analysis data completeness of the absolute number of COVID 19 deaths reported to the Integrated Disease Surveillance Programme (IDSP) on November 16 2020, and comparing them with the State-wise bulletins. We then identified seven states and one union territory based on the completeness and validity of reported COVID-19 deaths. ^8^

We extracted COVID-19 deaths for the age groups from IDSP Covid-19 special surveillance portal till the period of November 16 2020, and the daily COVID 19 bulletin of the respective states. The state-based COVID-19 mortality figures for the specific age groups were extracted from portal data and assigned as observed deaths by age group.^9^

### Population Estimations

The population for the country in 2020 was estimated using the census population figures of 2011 and the decadal growth rate.^10^ We then derived the age group wise population of India based on the current age demographic proportions.^11^ We developed the age structure using a uniform distribution of 15 years (<15 years, 15– 29 years, 30–44 years, 45–-59 years, 60–74 years, ≥75 years).

### Crude Mortality Rate and Age-adjusted Mortality Rate

The crude mortality rate of India and selected states were calculated from the total number of COVID-19 deaths to residents in the specified geographic area divided by the total population for the same geographic area (for a specified time period, usually a calendar year) and multiplied by 100,000. We then calculated the age-adjusted mortality rates by multiplying the crude mortality rate with the standardized mortality rate.

### Age Standardized mortality rate

We estimated the age-standardized mortality rates of India by calculating the number of COVID-19 deaths for each age group. This was then divided by the age group population that we estimated; for per million population.

We used the indirect standardization method to calculate the standardized mortality ratio (SMR) by age group for the states. We computed the expected deaths due to COVID-19 for each age group in each state. This was done by applying the estimated ASMR per million population of India (standard population) to the population figures of the state by specific age groups. We then calculated the SMR values for each state with 95% Confidence intervals. For estimating the age-standardized mortality rates, we multiplied the crude mortality rate of India with SMR for each state.

### The ratio of Age Standardised Mortality

In addition, we attempted to compare the mortality among the age groups within the selected states. We considered the age-standardized mortality of <15 year age group (as this age group was uniformly recorded with the lowest mortality) as the reference. We calculated the ratios of age-standardized mortality for each age group.

We performed all the analysis using Microsoft Excel 2018 and Open Epi (an open-source epidemiologic statistics for public health) Version 3.01. We summarised all the estimates in tables and compared them to bring out the state-wise differences in mortality.

### Ethical consideration

The study examined aggregate secondary data from COVID 19 web-based portal on death outcomes and age for selected states in the study period. This study protocol was approved by the public health emergency operational research review board of National Centre for Disease Control to initiate this analysis. Names and identifiers were delinked for confidentiality, and only an anonymized dataset was provided to the group on select variables for analysis.

## Results

### COVID 19 Deaths

India had reported a total of 118,883 deaths from March till November 2020. The COVID -19 deaths reported in the selected states by decreasing order were Maharashtra (35978, 30.3%), Delhi (5217, 4.4%), Karnataka (11541, 9.7%), Tamil Nadu (11440, 9.6%), Haryana (1960, 1.6%), West Bengal (5571, 4.7%), and Kerala (1888, 1.6%).

### Crude Mortality Rate and Age-adjusted Mortality Rate

The crude mortality rate of India stands at 88.9 per million population(118,883/1,337,328,910). The crude mortality rate (per million) was observed as highest for Maharashtra (270.4) and lowest in Kerala (53.5). However, the age-adjusted mortality rate (per million) was highest for Delhi (300.5) and lowest for Kerala (35.9). (Table II)

### Age Standardized Mortality Rate

The age standardized mortality rate (per million) for India was (<15 years=1.6, 15-29 years=6.3, 30-44 years=35.9, 45-59 years=198.8, 60-74 years =571.2, ≥75 years =931.6) (Table I).

**Table I.** **Age Standardized Mortality Rates for COVID 19 (per million population) in India and its selected states, 2020**

| Age(in years) | <15 | 15–29 | 30–44 | 45–59 | 60–74 | ≥75 |
| --- | --- | --- | --- | --- | --- | --- |
| Maharashtra | 6.5 | 14.5 | 98.1 | 522.1 | 1640.1 | 2818.1 |
| Delhi | 9.7 | 33.9 | 119.3 | 573.6 | 1937.3 | 3787.9 |
| Karnataka | 2.4 | 10.8 | 66.5 | 364.8 | 1111.8 | 2064.9 |
| Tamil Nadu | 2.2 | 7.0 | 45.4 | 231.7 | 773.6 | 1385.6 |
| Haryana | 2.1 | 4.5 | 31.6 | 146.4 | 489.3 | 837.5 |
| West Bengal | 1.0 | 4.8 | 18.1 | 102.3 | 346.1 | 748.4 |
| Kerala | 0.9 | 3.3 | 10.9 | 58.9 | 242.3 | 490.3 |
| <u>India</u> | 1.6 | 6.3 | 35.5 | 198.8 | 571.3 | 931.6 |

The age-standardized mortality rates (per million) in the selected states are summarised in Table II. Across all states, the highest mortality was observed in the age group of ≥75 years; it ranged from 490.3 (Kerala) to 3787.9 (Delhi). We also observed that the mortality rates ranged from 58.9 (Kerala) – 573.6 (Delhi) in the age group 45-59 years. The mortality rates were significantly lower in younger age groups (<29 years) across all states. [Table II.]

**Table II.** **Comparison of Crude and Age-Adjusted Mortality Rates for COVID-19, India and its selected States, 2020**

| India and its States | Crude Mortality Rate per million | Age-Adjusted Mortality Rate per million |
| --- | --- | --- |
| Maharashtra | 270.4 | 249.9 |
| **Delhi** | 259 | **300.5** |
| Karnataka | 175.4 | 172.4 |
| Tamil Nadu | 152.1 | 116.7 |
| Haryana | 67.8 | 73.7 |
| West Bengal | 57.6 | 53.8 |
| Kerala | 53.5 | 35.9 |
| **India** | 88.9 | - |

### The ratio of Age Standardised Mortality

While looking at the ratios for age-standardized mortality, we observed an increasing trend with age across the states. We observed that the COVID 19 mortality among 30-44 years compared to reference differed by a factor ranging between 12-28, while among the 45-59 year age group, the factor ranged between 59-152, and above 75 year age group the factor ranged between 390-860. [Figure 1.]

**Figure 1.**
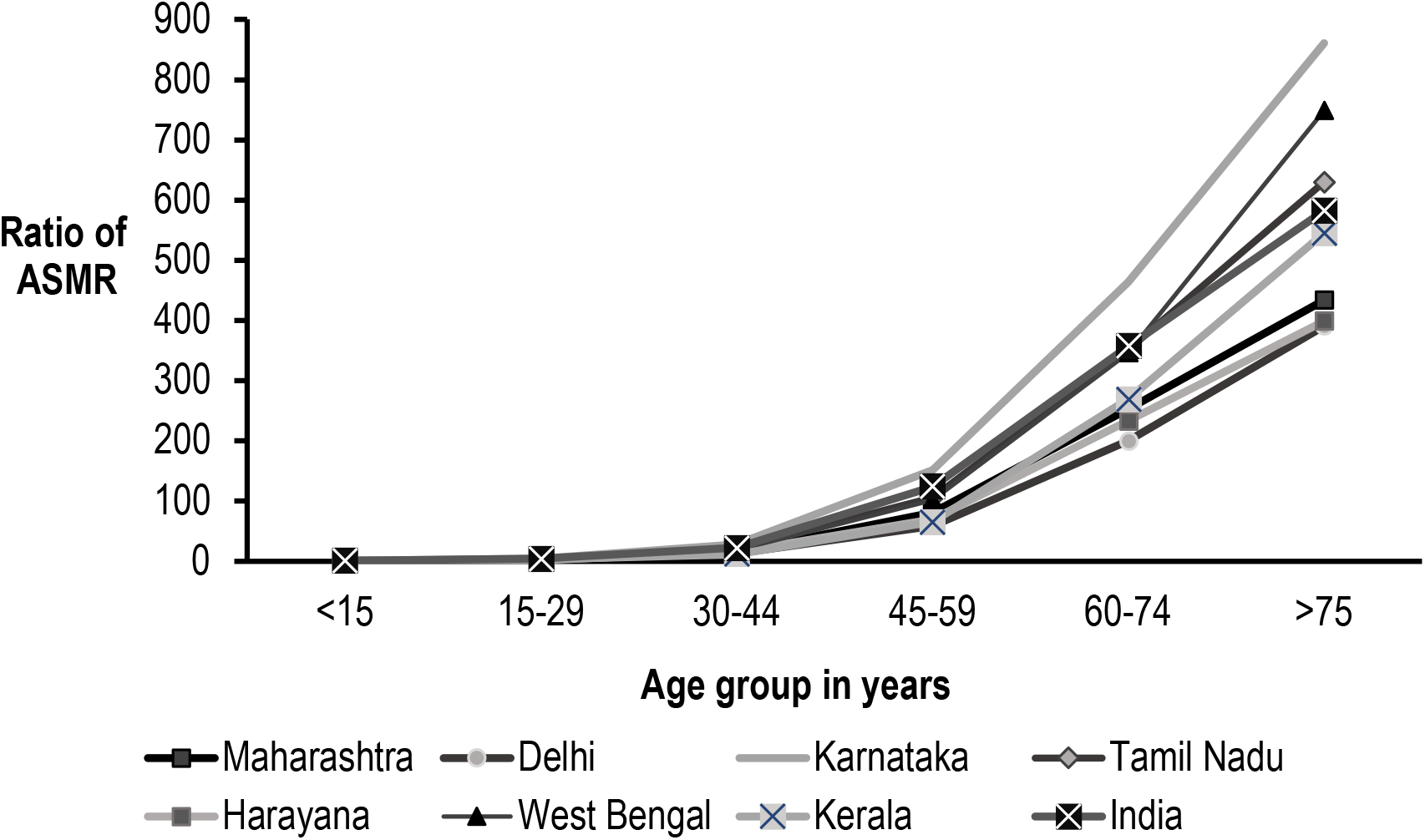
**Ratio of age-standardized mortality due to COVID-19 in India and its selected States, 2020**

## Discussion

Our study examined crude mortality rate, age-adjusted mortality rate, and age-standardized mortalities for COVID-19 in India and seven states. We observed a difference in the crude mortality rate when adjusted for age. Maharashtra, which seemed to have a high crude mortality rates, was replaced by Delhi on adjusting for age, while Kerala remained the lowest in both calculations. Standardization for age identified that the highest mortality rate was among the above 75 year age group; however, it also brought to notice the mortality rate of the 45-59 years age group. This finding was strengthened when comparing the age-standardized mortality within the states. This age group showed an increase in covid 19 mortality by more than sixty times the reference group.

The aggregate confirmed mortality rates tends to conceal the differential impact of the pandemic related to and levels of mortality by age due to demographic composition.^12^ Our findings demonstrate the importance of accounting for age when comparing rates in general and mortality in particular in Indian states. The population age structures are crucial for understanding those at highest risk of mortality both across and within countries for targeting policies to slow down transmission as well as intergenerational interactions.^13^ The overall mortality in India (89.9 deaths per million population) appears to be low in comparison to the world average; however, this may be attributed to India’s age distribution and also the data paucity in mortality figures.^14,15^ We would like to submit that the states with lower age-adjusted mortalities need to ensure the reported mortality figures are.

The findings of our analysis identify that the COVID-19 mortality risk is higher for the elderly population similar to other countries.^7, 16, 17^ However; we also observed that the mortality within the age group of 45-59 years which is an active working population group, has been sizeably affected across the states. This evidence is in concurrence with previous reports that suggest a higher caseload exists among the working population when compared to the population share of India.^14^ Thus it is imperative for the government to prioritize this age group in their targeted interventions, implement and ensure adherence to mitigation behaviours and completion of vaccination.

We faced limitations in our analysis with regard to the provisional nature of mortality data extracted from the Integrated Disease Surveillance Programme special surveillance portal up to November 16, 202 0. We tried to validate the mortality data with daily COVID 19 bulletins released by the states to ensure greater accuracy; some inconsistencies with the final mortality numbers cannot be ruled out. However, they are unlikely to impact the estimate and standardized mortality rates for comparison between the states and age groups.

## Conclusion

We conclude that standardized mortality rates are the need of the hour to identify the vulnerable states and vulnerable age group for public health action. We have reported evidence on the high COVID 19 mortality among the elderly age group and also identified the impact of COVID 19 on the working population. We recommend further workup and evaluation of age-standardized mortality estimates in all the states and also the inclusion of other variables like gender, socio-economic status for standardization while identifying at-risk populations and implementing targeted evidence-based public health actions.

## Data Availability

THE DATA WAS EXTRACTED FROM COVID 19 SPECIAL SURVEILLANCE PORTAL IMPLEMENTED BY NATIONAL CENTRE FOR DISEASE CONTROL (NCDC).

## Acknowledgement

We acknowledge Dr Naman K Shah, Adjunct Professor, ICMR-National Institute of Epidemiology, for his valuable contribution in reviewing the manuscript and providing suggestions.

## Financial support and Sponsorship

No funding support was received

## Conflicts of Interest

There are no conflicts of interest

---

**(Supplementary) Table III:** **Comparison of Standardized Mortality Ratios of the specific States in India, 2020**

| India and its States | **Standardized Mortality Ratio** | **(95% CI)** |
| --- | --- | --- |
| **Maharashtra** | 2.8 | (2.8-2.8) |
| **Delhi** | 3.4 | (3.3-3.5) |
| **Karnataka** | 1.9 | (1.9-2.0) |
| **Tamil Nadu** | 1.3 | (1.3-1.3) |
| Haryana | 0.8 | (0.8-0.9) |
| West Bengal | 0.6 | (0.6-0.6) |
| Kerala | 0.4 | (0.4-0.4) |

## Notes

### Competing Interest Statement

The authors have declared no competing interest.

## References

1. ECDC. COVID-19 situation update worldwide, as of July 13 2020. Situation updates worldwide 2020 [Internet]. Available from: https://www.ecdc.europa.eu/en/geographical-distribution-2019-ncov-cases. Accessed July 13 2020.

2. R, Lavergne, and McGrail K. ‘Amenable (or Avoidable) Mortality as an Indicator of Health System Effectiveness.’ Healthcare Policy = Politiques de Sante 10, no. 1 (January 1 2014): 8–9.

3. ‘WHO | SDG 3: Ensure Healthy Lives and Promote Wellbeing for All at All Ages. World Health Organization. Accessed April 29 2020. http://www.who.int/sdg/targets/en/.

4. ECDC. ‘COVID-19 Situation Update Worldwide, as of November 17 2020’ [Internet]. https://www.ecdc.europa.eu/en/geographical-distribution-2019-ncov-cases. Accessed November 18 2020.

5. Stawicki SP, Jeanmonod R, Miller AC, Paladino L, Gaieski DF, Yaffee AQ et al. The 2019-2020 Novel Coronavirus (Severe Acute Respiratory Syndrome Coronavirus 2) Pandemic: A Joint American College of Academic International Medicine-World Academic Council of Emergency Medicine Multidisciplinary COVID-19 Working Group Consensus Paper. J Glob Infect Dis. 2020; 12(2):47–93.

6. Ram U, Jha P, Gerland P, Hum RJ, Rodriguez P, Suraweera W, et al. Age-specific and sex-specific adult mortality risk in India in 2014: analysis of 0·27 million nationally surveyed deaths and demographic estimates from 597 districts. The Lancet Global Health. 2015 Dec;3(12):e767–75.

7. Goldstein JR, Lee RD. Demographic perspectives on the mortality of COVID-19 and other epidemics [published correction appears in Proc Natl Acad Sci U S A. 2020; 117(47):29991]. Proc Natl Acad Sci U S A. 2020; 117(36):22035–41.

8. Government of India. COVID-19 State-wise Status from Ministry of Health and Family Welfare [Internet]. Available from: https://www.mohfw.gov.in/ Accessed November 18 2020.

9. Government of India. COVID-19 surveillance portal [Internet]. Available from: https://covid19.nhp.gov.in/ Accessed November 16 2020

10. Population projections for India and states 2011 – 2036’. National Commission on population ministry of health & family welfare Nirman Bhawan, New Delhi – 110011, November 2019. Available from: https://nhm.gov.in/New_Updates_2018/Report_Population_Projection_2019.pdf Accessed November 18 2020

11. Office of the Registrar General & Census Commissioner, India SRS Statistical Report 2018. Available from: https://censusindia.gov.in/vital_statistics/SRS_Reports_2018.html Accessed November 18 2020

12. Joy M, Hobbs FR, Bernal JL, Sherlock J, Amirthalingam G, McGagh D, Akinyemi O, Byford R, Dabrera G, Dorward J, Ellis J. Excess mortality in the first COVID pandemic peak: cross-sectional analyses of the impact of age, sex, ethnicity, household size, and long-term conditions in people of known SARS-CoV-2 status in England. British Journal of General Practice. 2020 Dec 1;70(701):e890–8.

13. Dowd J, Andriano L., Brazel D., Rotondi V., Block P., Ding X. et al. Demographic science aids in understanding the spread and fatality rates of COVID-19. Proc Natl Acad Sci U S A. 2020; 117(18): 9696–98.

14. Philip M., Ray D., Subramanian S. Decoding India’s Low Covid-19 Case Fatality rate [Internet]. Working paper 27696, August 2020. Accessed November 16 2020

15. Zimmermann L, Salvatore M, Babu G, Mukherjee B. Estimating COVID-19 Related Mortality in India: An Epidemiological Challenge with Insufficient Data.

16. Hallal PC. Worldwide differences in COVID-19-related mortality. Cien Saude Colet. 2020; 25(suppl 1): 2403–10.

17. Mathers, Colin D., and Dejan Loncar. ‘Projections of Global Mortality and Burden of Disease from 2002 to 2030’. PLOS Med. 2006; 3(11): e442. https://doi.org/10.1371/journal.pmed.0030442.

